# Catastrophic Hunger in Gaza: Unprecedented Levels of Hunger Post-October 7^th^. A Real Population-Based Study from the Gaza Strip

**DOI:** 10.1101/2024.08.21.24312386

**Authors:** MoezAlIslam Faris, Ayman S. Abutair, Reham M. Elfarra, Nida. A. Barqawi, Amal M. Firwana, Rawan M. Firwana, Madleen M. AbuHajjaj, Shaimaa A. Shamaly, Samar S. AbuSamra, Hanan S. Bashir, Noor A. Abedalrahim, Noor A. Nofal, Mhran K. Alshawaf, Rania M. Al Shatali, Kafa I. Ghaben, Moayad I. Alron, Sara S. Alqeeq, Aya O. Al-Nabahin, Reem A. Badawi

## Abstract

**Background:** The Gaza Strip, spanning approximately 365 square kilometers, has been a focal point of geopolitical tensions and humanitarian crises. The military escalation on October 7^th^ exacerbated existing vulnerabilities, notably food security and hunger, with an estimated 85,750 deaths due to Israeli attacks, representing about 8% of the 2.34 million population. This research aims to inform policymakers and humanitarian organizations about strategies to mitigate hunger and improve food security in Gaza amidst this damaging war.

**Methods:** A cross-sectional study was conducted from May to July 2024, assessing food insecurity and hunger among Palestinian households across the five governorates of Gaza. The study applied a quantitative research approach; the study involved 1209 households. The study utilized the Household Food Security Survey Module (HFSSM), Household Food Insecurity Access Scale (HFIAS), and Household Hunger Scale (HHS) to measure food insecurity and hunger. Self-reported anthropometric data were also collected. Data were analyzed using SPSS version 29, employing correlation tests, chi-square analysis, and logistic regression.

**Results:** Out of 1503 surveyed households, 1209 were included in the final analysis. The mean age of participants was 38 years, with 53.5% being female. Significant findings included high levels of food insecurity, with 100% of households experiencing some level of food insecurity according to HFSSM and 97.7% being severely food insecure per HFIAS. The results revealed a catastrophic, unprecedented, extremely high prevalence of hunger, reaching more than 95%. The HHS indicated that 88% of households experienced varying degrees of hunger. The war was associated with significant weight loss among individuals, with the average weight dropping from 74.6 kg before the war to 64.1 kg. Factors such as displacement, age, economic status, and educational level significantly impacted hunger severity.

**Conclusion:** The study reveals a severe food insecurity and hunger crisis in the Gaza Strip, exacerbated by the ongoing damaging war by Israeli forces. The findings highlight the urgent need for coordinated efforts to address food security and support the resilience and recovery of affected communities. Policymakers and humanitarian organizations must develop targeted interventions to mitigate hunger and improve the living conditions in Gaza.

## Introduction

The Gaza Strip, a small territory of approximately 365 square kilometers, has long been a focal point of geopolitical clashes, socioeconomic challenges, and humanitarian crises. Following the escalation of military actions on October 7^th^, the region has experienced heightened levels of Israeli forces attacks, resulting in significant disruptions to infrastructure, everyday life, and essential services. This has further exacerbated existing vulnerabilities, particularly concerning food security and the prevalence of hunger among the Gazanian population. As a sequela, it has been estimated that about 85,750 deaths resulted only in the Gaza Strip caused by Israeli attacks, accounting for about 8% of the 2.34 million population [1]. This horrible killing rate emphasizes the urgent need for a ceasefire and humanitarian aid, highlighting the critical importance of documenting the full extent of the tragedy for historical accountability and future recovery efforts.

Hunger and food insecurity are critical issues that impact the human health, well-being, and socioeconomic stability of communities [2]. In conflict zones like Gaza, these issues are often intensified due to factors such as restricted access to food supplies, destruction of agricultural resources, displacement of populations, and the breakdown of local markets. The interplay between ongoing political conflict and food insecurity necessitates a comprehensive assessment to understand the full extent and implications of hunger in the region [3]. The period following October 7^th^ has seen numerous reports of severe shortages in food availability, restricted access to essential goods, and a significant rise in the number of individuals and families experiencing hunger [4,5]. These developments underline the urgency for empirical research to quantify and analyze the prevalence of hunger and famine in Gaza, providing a basis for targeted interventions and policy responses.

The Household Hunger Scale (HHS) is a significant tool in assessing the prevalence of famine, hunger, and food insecurity. Developed by the Food and Agriculture Organization (FAO) and the United States Agency for International Development (USAID), the HHS measures the degree of hunger within a household based on a set of standardized questions, providing a direct assessment of hunger by focusing on experiences and behaviors associated with food scarcity [6]. Its standardized set of questions ensures consistency in data collection and comparability across different regions and populations, allowing for reliable tracking of hunger trends. The HHS identifies and categorizes the severity of hunger, distinguishing between mild, moderate, and severe hunger, thus offering nuanced insights into food insecurity. By focusing on households, the HHS captures the micro-level impacts of food insecurity, revealing how individual families cope with food shortages, which is critical for understanding the immediate effects of food crises. The data from the HHS informs policymakers and humanitarian organizations about the most affected areas and populations, aiding in the design of targeted interventions and effective resource allocation [7].

Additionally, HHS serves as a tool for monitoring and evaluating the effectiveness of food security programs, tracking changes in household hunger over time to assess the impact of interventions. Acting as an early warning system, the HHS can detect signs of increasing hunger before they escalate into famine, allowing for timely responses and preventive measures. In crises, it quickly assesses the extent of hunger and food insecurity, which is crucial for emergency response and relief efforts. The HHS methodology often involves local communities in data collection, fostering a sense of ownership and empowerment, ensuring data accuracy and relevance, and supporting advocacy efforts by providing concrete evidence of hunger to raise awareness and mobilize support for food security initiatives. Thus, the HHS is indispensable for policymakers, humanitarian organizations, and communities in addressing and mitigating food crises. In an era of extreme scarcity of humanitarian funding, HHS has substantial implications for resource allocation and humanitarian prioritization [8]. The Household Food Insecurity Access Scale (HFIAS) and the Household Food Security Survey Module (HFSSM) are also pivotal yet contested tools in assessing food security and hunger.

The findings of this research are intended to inform policymakers, humanitarian organizations, and international bodies, facilitating the development of effective strategies to mitigate hunger and enhance food security in the occupied Strip amidst the ongoing unequalized, damaging war. Through this assessment, we aim to highlight the urgent need for coordinated efforts to address the humanitarian crisis and support the resilience and recovery of the affected communities in the Gaza Strip.

## Methods

### Study settings, population, and tools

A cross-sectional design was followed in assessing the prevalence of food insecurity and hunger in the Gaza Strip from May to July 2024, after seven to nine months of the Israeli military attacks in response to the 7^th^ October attack by Hamas. The study applied quantitative research methodology, using a non-probability convenience sampling technique, covering the Palestinian households in the five governorates of the Gaza Strip that are the northern governorates (North Gaza, Gaza City), the middle (Deir Al Balah), and the southern (Khan Younis, Rafah).

The Palestinian Ministry of Health in the Gaza Strip granted ethical approval. Informed consent was obtained from all participants after the study’s aims, advantages, risks, information confidentiality, and voluntary nature of participation were discussed. Furthermore, all data collectors and investigators ensured the confidentiality of the information gathered from each study participant by using code numbers instead of personal identifiers and making the questionnaire inaccessible to anyone other than the investigators.

For sample size calculation, we used the single population proportion formula by using Epi Info StatCalc considering the following assumptions: 95% confidence level (Zα/2), 32.4% proportion of households fell into the household hunger categories [9] for prevalence (p), and 5% margin of error (d), which was 337. With a 30% non-response rate and a three-size effect, the calculated sample size was 1213 households. A non-probability convenience sampling technique was used to select the required sample of households from all the affected parts of the Gaza Strip.

### Food insecurity and hunger assessment and scoring systems

#### The HFSSM

The HFSSM tool consists of 18 questions; 11 of them are yes/no questions, and 7 of them have a 4-point Likert scale from never to often, where never and rarely are considered zero scores and sometimes given one mark score. The maximum total score was 18 points, and then it was categorized into two categories, with a score of 1 or more being enough to indicate food insecurity. So, only the participants who got a score of zero indicated food security [10,11].

#### The HFIAS

HFIAS tool consists of 9 questions using a 4-point Likert scale (never=0, rarely=1, sometimes=2, often=3). The maximum total score was 27 points, then this total score was categorized into four categories as follows: 1 = Food Secure if [(Q1=0 or Q1=1) and Q2=0 and Q3=0 and Q4=0 and Q5=0 and Q6=0 and Q7=0 and Q8=0 and Q9=0], 2=Mildly Food Insecure Access if [Q1=2 or Q1=3 or Q2=1 or Q2=2 or Q2=3 or Q3=1 or Q4=1) and Q5=0 and Q6=0 and Q7=0 and Q8=0 and Q9=0], 3=Moderately Food Insecure Access if [(Q3=2 or Q3=3 or Q4=2 or Q4=3 or Q5=1 or Q5=2 or Q6=1 or Q6=2) and Q7=0 and Q8=0 and Q9=0], and 4=Severely Food Insecure Access if [Q5=3 or Q6=3 or Q7=1 or Q7=2 or Q7=3 or Q8=1 or Q8=2 or Q8=3 or Q9=1 or Q9=2 or Q9=3] [12].

#### The HHS

The HHS consists of three simple questions about the experience of extreme hunger with a yes/no answer, and if the answer is yes, there is a follow-up question about the frequency. The recall period is the previous 30 days. The HHS tool contains only three questions 4-point Likert scale (never =0, rarely, and sometimes= 1, often=2). The total possible score was 6 points. The cutoff points were as follows: 0-1; little or no household hunger; 2-3; moderate household hunger; 4-6; severe household hunger [6].

### Self-reported anthropometric measurements

For this study, the three assessment tools were translated into Arabic by expert in the two languages. To ensure parallel form reliability, the Arabic versions of the questionnaires were back-translated into English. The original and back-translated English versions of the questionnaires were compared for accuracy. Additionally, self-reported anthropometric measurements, comprising height and weight, were questioned before and during the war. These measurements were used to calculate and categorize the body mass index (BMI, kg/m^2^). BMI categories, based on the definition of the WHO, include underweight (BMI < 18.5), normal weight (BMI 18.5 - 24.9), overweight (BMI 25 - 29.9), and obesity (BMI ≥ 30 kg/m^2^), enabling further insight into participants’ weight status [13].

### Statistical analysis

Data were analyzed using Statistical Package for Social Science (SPSS) software, version 29 (IBM Corp, Armonk, NY, USA). Categorical variables were reported as frequencies and percentages, whereas continuous variables were described as mean ± standard deviation (SD). All of the scale questions were expressed as frequency and percentages of the responses among participants. The total score was calculated as mentioned above in the scoring paragraph, and the cutoff points were applied to categorize the total score into its categories. The correlation between the total score of the HFSSM, HFIAS, and HHS scales and the sociodemographic data was determined by using the correlation test. Cross-tabulation and Chi-square analysis were used to investigate the categories of the HHS scale among sociodemographic variables. A logistic regression analysis determined significant predictors of suffering from hunger. A confidence interval (CI) of 95% was applied to represent the statistical significance of the results, and the level of significance was predetermined as P < 0.05.

## Results

One thousand five hundred-three households from the five governorates were surveyed. After data cleaning and removing participants with missing data, 1209 households were included in the final statistical analysis. These 1209 households were presented by 1209 representatives (aged 38 ± 9.6 years, 53.5% females). Households were primarily from the Middle region (Deir Al Balah), which accounts for 65.0%. The Northern region (North Gaza and Gaza City) accounted for 27%, while the Southern region (Khan Younis and Rafah) represented about 8%.

Most households were headed by men (about 90%), with an average of 5.63 ± 1.79 individuals per family household and 3.09 ± 1.59 dependent children. Before the war, 34% of the participants were of low economic status, 58% were of medium status, and about one-fifth were living in camps while the rest were living in city houses. A vast majority of the participants were married (96.0%), with small percentages being widowed or divorced. Among partners (38.36 ± 10.28, range 16-95 years), 62.2% were not working, and the most common educational level was a Bachelor’s degree (49.0%). The Israeli attacks caused complete house destruction for 54.3% of the households and partial destruction for 30.8%, with about 59% currently living in tents and 24% in schools. About 78% received intermittent help from relief organizations, 19.4% received no help, and 2.5% received regular help. During the 7-9 months of the war, the household families experienced 4.45 ± 2.49 displacements across the Strip regions (Table 1).

**Table 1:**
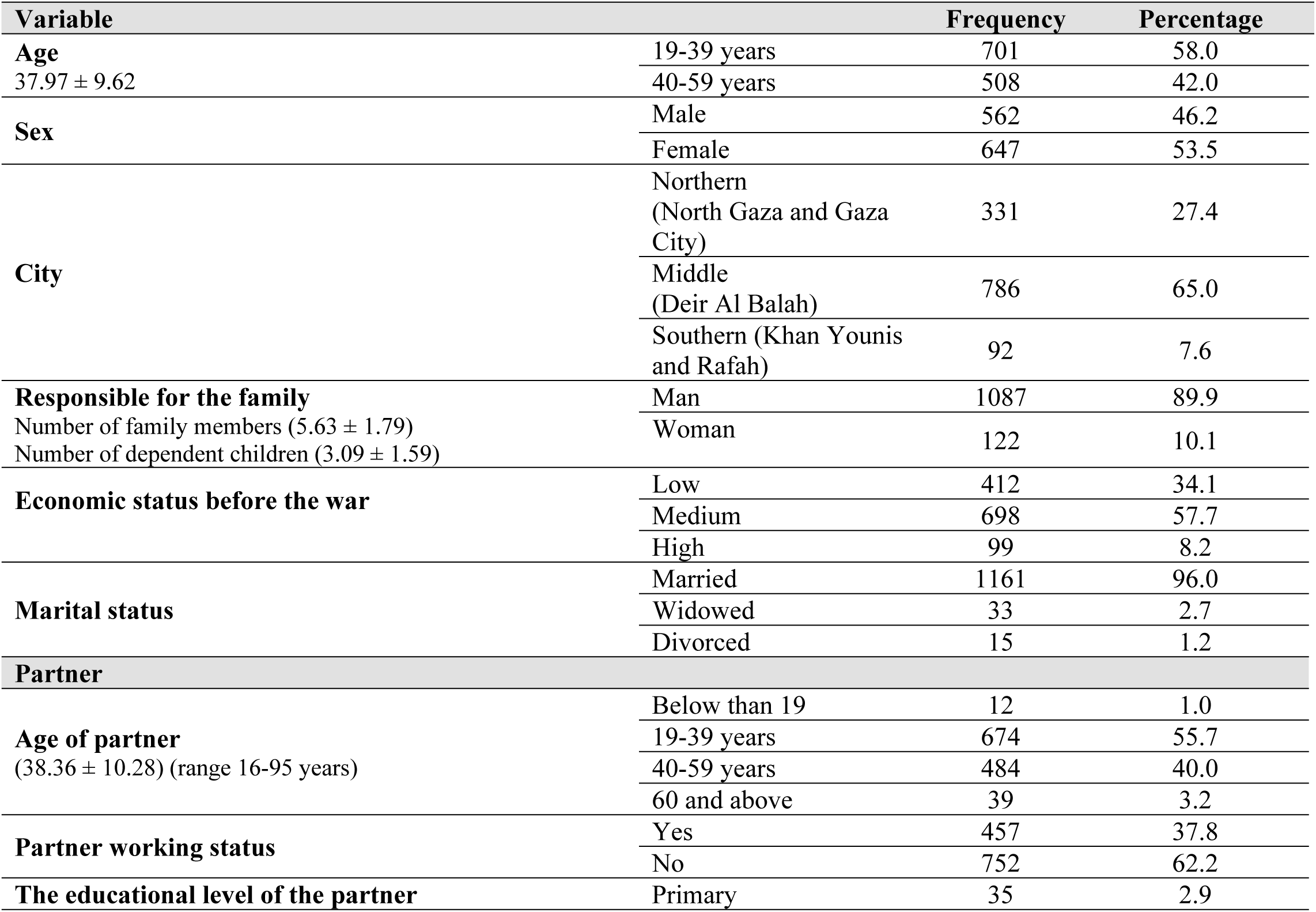
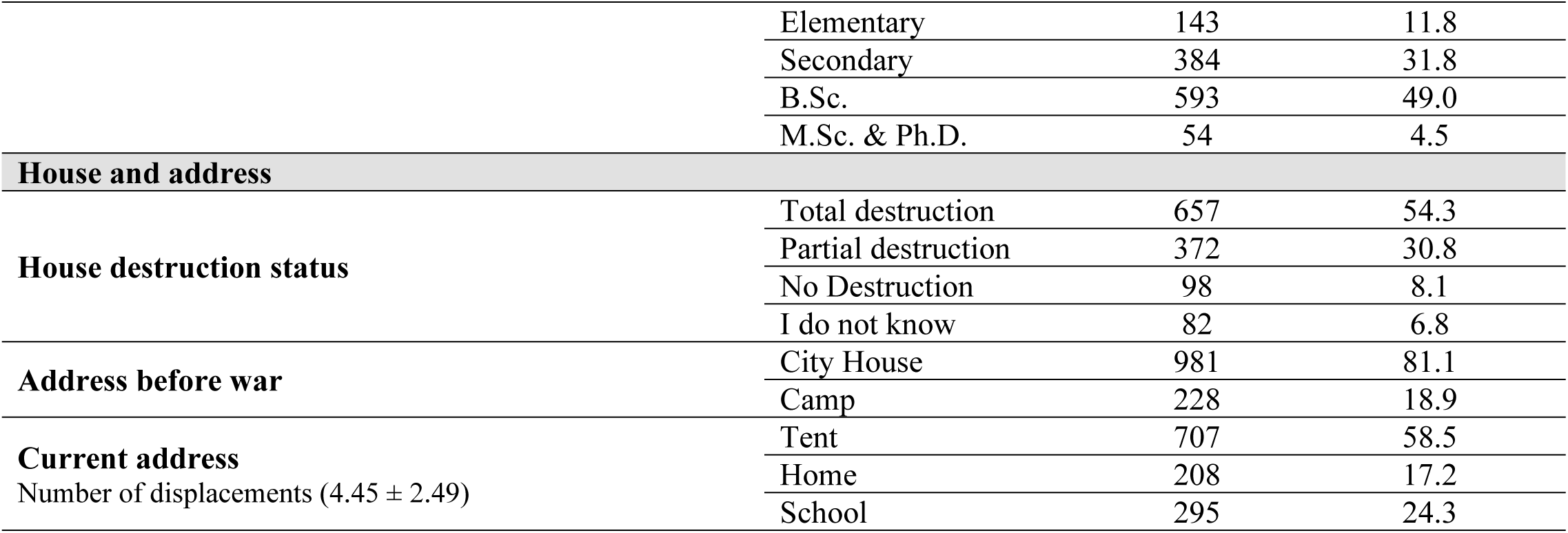
Sociodemographic characteristics of the study participants and their represented households (n=1209).

The results presented in Table 2 provide an overview of malnutrition and anthropometric characteristics of the 1209 households’ children. A significant majority, about 84% (n=1013) of the households’ children exhibited one or more of the starvation symptoms (significant weight loss, fatigue, weakness, irritability, and a decreased immune response), with an average number of children who have symptoms 1.91 ± 1.47 per household. Very few children, 0.4% (n=5) of households\ children passed away because of starvation, as reported by their parents, with average number of children who passed away because of starvation was 0.01 ± 0.17 per household.

**Table 2.** Malnutrition and anthropometric characteristics of the surveyed households (n=1209)

| Variable |  | Frequency | percentage |
| --- | --- | --- | --- |
| **Starvation symptoms appearance**<br>The number of children who have symptoms ($1.91 \pm 1.47$) | Yes | 1013 | 83.8 |
|  | No | 196 | 16.2 |
| **Did any child die because of starvation?**<br>Number of children who die because of starvation ($0.01 \pm 0.17$) | Yes | 5 | 0.4 |
|  | No | 1204 | 99.6 |
| **Do you receive any help from a relief organization?** | No | 235 | 19.4 |
|  | Regularly | 30 | 2.5 |
|  | Intermittent | 944 | 78.1 |
| Variable |  | Mean | SD. |
| Weight before war (kg) |  | 74.61 | 15.91 |
| Height (cm) |  | 168.48 | 10.11 |
| Number of kilograms lost since war started (kg) |  | 10.52 | 8.48 |
| Current calculated weight (= weight before the war (kg)-number of kg lost since war started) |  | 64.09 | 15.23 |
| BMI (current), kg/m<sup>2</sup> |  | 22.60 | 5.16 |
| BMI (before the war), kg/m<sup>2</sup> |  | 26.30 | 5.36 |
| BMI (kg/m<sup>2</sup>) categories [n(%)] |  | Current | Before war |
| Underweight (18.5-24.9) |  | 188 (15.6) | 49 (4.1) |
| Normal (25-29.9) |  | 698 (57.7) | 461 (38.1) |
| Overweight (30-34.9) |  | 255 (21.1) | 447 (37.0) |
| Obese (>35) |  | 68 (5.6) | 252 (20.8) |

In terms of assistance from relief organizations, about one-fifth (19.4%, n=235) reported receiving no help, while the vast majority (78%, n=944) received intermittent assistance. Overall, the data indicates a high prevalence of starvation symptoms among children. Most families received intermittent assistance from relief organizations, with a notable proportion not receiving any help.

Before the war, the mean body weight was 74.6 ± 15.9 kg. On average, households’ individuals lost 10.5± 8.5 kg since the war started, resulting in a mean current calculated weight of 64.1±15.2 kg. The mean current BMI was 22.6±5.2 kg/m^2^, while the mean before the war was 26.3±5.4. In terms of BMI categories, the current distribution shows that about 16% of individuals were underweight, 58% had a normal weight, 21% were overweight, and 6% were obese. Before the war, these values were about 4%, 38%, 37%, and 21%, respectively. These results indicate a significant shift in the weight and BMI categories of the individuals, with a general trend of weight loss and a reduction in the proportion of overweight and obese individuals following the war (Table 2).

The HFSSM survey questions about experiences with food insecurity in the past year. Most respondents (over 50%) reported worrying about running out of food before they had money to buy more, did not have money to get more food, and more than two-thirds were not able to afford balanced meals.

For questions specifically about adults, nearly all respondents (over 95%) in the households cut meal sizes or skipped meals at some point in the last year and, particularly, in three or more months, and reported that they ate less than they feel should do. About 80% of interviewed household adults experienced hunger but did not eat, and over 90% reported that they lost body weight. Less than half (43%) of respondent household adults went a whole day without eating at some point in the last year, with particular attention toward the three or more months by about one-third of the respondents.

For questions concerning children, the most common response (about two-thirds) was that children relied on a few cheap foods, and their parents could not feed them balanced meals. Nearly as many respondents (over 55%) said their children were not eating enough, and over 80% reported having to cut their children’s meal sizes or described their children being hungry or having their children skip meals, especially during three or more months (about 80%). Half of the respondents said their children went a whole day without eating at some point in the last year. The survey concludes that 100% of the sample was food insecure (Table 3).

**Table 3:** HFSSM questionnaire for food insecurity (18 items)

| Questions |  |  | Never | Rarely | Sometimes | Often |
| --- | --- | --- | --- | --- | --- | --- |
| Household | Q1 | Worry that food would run out before (I/we) got money to buy more | 67 (5.5) | 82 (6.8) | 428 (35.4) | 632 (52.3) |
| Household | Q2 | The food bought did not last, and (I/we) did not have money to get more | 43 (3.6) | 138 (11.4) | 396 (32.8) | 632 (52.3) |
| Household | Q3 | Could not afford to eat balanced meals | 47 (3.9) | 55 (4.5) | 276 (22.8) | 831 (68.7) |
|  |  |  | Yes |  | No |  |
| Adult | Q4 | Adult(s) cut the size of meals or skipped meals | 1175 (97.2) |  | 34 (2.8) |  |
| Adult | Q5 | Adult(s) cut the size of meals or skipped meals in three or more months | 1150 (95.1) |  | 59 (4.9) |  |
| Adult | Q6 | Respondent ate less than felt he/she should | 1171 (96.9) |  | 38 (3.1) |  |
| Adult | Q7 | The respondent was hungry but did not eat | 944 (78.1) |  | 265 (21.9) |  |
| Adult | Q8 | Respondent lost weight | 1118 (92.5) |  | 91 (7.5) |  |
| Adult | Q9 | Adult(s) did not eat for the whole day | 523 (43.3) |  | 686 (56.7) |  |
| Adult | Q10 | Adult(s) did not eat for a whole day in three or more months | 393 (32.5) |  | 816 (67.5) |  |
|  |  |  | Never | Rarely | Sometimes | Often |
| Child | Q11 | Relied on few kinds of low-cost food to feed the child(ren) | 44 (3.6) | 66 (5.5) | 282 (23.3) | 817 (67.6) |
| Child | Q12 | Could not feed the child(ren) balanced meals | 25 (2.1) | 72 (6.0) | 315 (26.1) | 797 (65.9) |
| Child | Q13 | The child (ren) was not eating enough | 43 (3.6) | 105 (8.7) | 379 (31.3) | 682 (56.4) |
|  |  |  | Yes |  | No |  |
| Child | Q14 | Cut size of child(ren's) meals | 1032 (85.4) |  | 177 (14.6) |  |
| Child | Q15 | Child(ren) were hungry | 1017 (84.1) |  | 192 (15.9) |  |
| Child | Q16 | Child(ren) skipped meals | 1033 (85.4) |  | 176 (14.6) |  |
| Child | Q17 | Child(ren) skipped meals in three or more months | 960 (79.4) |  | 249 (20.6) |  |
| Child | Q18 | The child (ren) did not eat for the whole day | 598 (49.5) |  | 611 (50.5) |  |

**The total HFSSM score was $14.54 \pm 2.83$ (range 3.0 – 18.0), and 100% of the sample size is considered to be food insecurity.**

Similar to the HFSSM survey, the HFIAS survey revealed that most respondents (almost 50%) reported worrying that their household would not have enough food. In terms of variety and quality of food, about 80% of respondents said they were not able to eat the preferred kinds of food, and almost 61% reported eating a limited variety of foods. Over 65% said they had to eat some foods they did not want to eat, and over half (56%) said they ate smaller meals than they felt they needed. The survey also asked about going a whole day and night without eating. Nearly one-third of respondents reported going to sleep hungry, while about one-fifth reported that they went a whole day and night without eating. The results show that almost all (98%) of the sample was severely food insecure, with 2.3% experiencing moderate food insecurity. No respondents were categorized as mildly food insecure. The total HFIAS score was 19.85 ± 4.54 (range 4.0 – 27.0), implying an extremely high level of food insecurity (Table 4).

**Table 4:** HFIAS questionnaire (9 items)

| Questions |  | Never | Rarely | Sometimes | Often |
| --- | --- | --- | --- | --- | --- |
| Q1 | Worry that the household would not have enough food | 68 (5.6) | 76 (6.3) | 481 (39.8) | 584 (48.3) |
| Q2 | Not able to eat the kinds of food preferred | 42 (3.5) | 39 (3.2) | 174 (14.4) | 954 (78.9) |
| Q3 | Eat a limited variety of foods | 112 (9.3) | 61 (5.0) | 303 (25.1) | 733 (60.6) |
| Q4 | Eat some foods that you really did not want to eat | 30 (2.5) | 85 (7.0) | 285 (23.6) | 809 (66.9) |
| Q5 | Eat a smaller meal than you feel you need | 67 (5.5) | 100 (8.3) | 364 (30.1) | 678 (56.1) |
| Q6 | Eat fewer meals in a day | 48 (4.0) | 115 (9.5) | 299 (24.7) | 747 (61.8) |
| Q7 | No food to eat of any kind in your household | 174 (14.4) | 254 (21.0) | 373 (30.9) | 408 (33.7) |
| Q8 | Go to sleep at night hungry | 148 (12.2) | 247 (20.4) | 428 (35.4) | 386 (31.9) |
| Q9 | Go a whole day and night without eating | 298 (24.6) | 352 (29.1) | 313 (25.9) | 246 (20.3) |

|  | Frequency | Percent |
| --- | --- | --- |
| **The total HFIAS score was $19.85 \pm 4.54$ (range 4.0 – 27.0)** |  |  |
| Severely food-insecure access | 1181 | 97.7 |
| Moderately food-insecure access | 28 | 2.3 |
| Mildly food-insecure access | 0 | 0.0 |
| Food Secure | 0 | 0.0 |

Table 5 shows responses to the HHS, which focuses specifically on going hungry in the surveyed households. Over a third (33.7%) of respondents reported having no food to eat at all at some point in the last 30 days. Similarly, over a third (35.4%) said they went to sleep hungry, and nearly one-fifth (20%) used to go a whole day and night without eating. The survey results indicate an extremely high prevalence of hunger within the sample. While 11.7% reported little to no household hunger, the vast majority (89%) experienced hunger in varying degrees, with about 41% experiencing moderate hunger and nearly half (48%) experiencing severe hunger in their household during the last 30 days.

**Table 5:** HSS questionnaire (3 items)

| **Questions** |  | **Never** | **Rarely** | **Sometimes** | **Often** |
| --- | --- | --- | --- | --- | --- |
| Q1 | No food to eat of any kind in your household | 174 (14.4) | 254 (21.0) | 373 (30.9) | 408 (33.7) |
| Q2 | Go to sleep at night hungry | 148 (12.2) | 247 (20.4) | 428 (35.4) | 386 (31.9) |
| Q3 | Go a whole day and night without eating | 298 (24.6) | 352 (29.1) | 313 (25.9) | 246 (20.3) |

|  |  | **Frequency** | **Percent** |
| --- | --- | --- | --- |
| **The total HHS score was $3.35 \pm 1.44$ (range 0.0 – 6.0)** |  |  |  |
|  | Little or no household hunger | 141 | 11.7 |
|  | Moderate household hunger | 493 | 40.8 |
|  | Severe household hunger | 575 | 47.6 |

The correlation Table 6 examines the relationship between three measures of hunger and food insecurity (HHS, HFSSM, HFIAS) and various sociodemographic determinants. There was a weak but statistically significant positive correlation between the number of times a household was displaced and all three hunger/food insecurity scores. This means that more displacement was associated with higher hunger and food insecurity scores. For the age factor, **t**here were weak but statistically significant positive correlations between age and all three hunger/food insecurity scores. This suggests that older individuals may be more likely, but not necessarily, to experience hunger or food insecurity. The age of the partner also shows a similar correlation with HHS and HFIAS scores. For children’s health**, t**he number of children with symptoms shows a weak but statistically significant positive correlation with all three hunger/food insecurity scores, implying a link between children’s health and household hunger and that the food security status of the household directly impacts children.

**Table 6:** Correlation between the three food security/hunger assessment tools and the sociodemographic characteristics of the study households in the Gaza Strip.

| Sociodemographic characteristics |  | HHS total score | HFSSM total score | HFIAS total score |
| --- | --- | --- | --- | --- |
| Number of displacement times | Pearson Correlation | **0.161\*\*** | **0.077\*\*** | 0.036 |
|  | P-value | **0.000** | **0.008** | 0.208 |
| Age | Pearson Correlation | **0.139\*\*** | **0.090\*\*** | **0.098\*\*** |
|  | P-value | **0.0001** | **0.002** | **0.001** |
| Age of partner | Pearson Correlation | **0.132\*\*** | 0.046 | **0.106\*\*** |
|  | P-value | **0.0001** | 0.112 | **0.0001** |
| Number of family members | Pearson Correlation | 0.052 | 0.023 | 0.005 |
|  | P-value | 0.069 | 0.420 | 0.864 |
| Number of dependent children | Pearson Correlation | 0.054 | -0.001 | 0.022 |
|  | P-value | 0.063 | 0.985 | 0.435 |
| How many children have symptoms | Pearson Correlation | **0.150\*\*** | **0.092\*\*** | **0.071\*** |
|  | P-value | **0.0001** | **0.001** | **0.014** |
| How many children died because of starvation | Pearson Correlation | 0.036 | **0.069\*** | 0.036 |
|  | P-value | 0.209 | **0.017\*** | 0.215 |
| Weight before war | Pearson Correlation | -0.052 | -0.014 | -0.009 |
|  | P-value | 0.073 | 0.616 | 0.767 |
| How many kilograms have you lost from weight since the war started | Pearson Correlation | 0.039 | -0.048 | **-0.082\*\*** |
|  | P-value | 0.180 | 0.098 | **0.004** |
| Current calculated weight | Pearson Correlation | **-.075\*\*** | 0.011 | 0.037 |
|  | P-value | **0.009** | 0.691 | 0.203 |
| BMI current | Pearson Correlation | **-.097\*\*** | -0.044 | 0.006 |
|  | P-value | **0.001** | 0.128 | 0.834 |
| BMI before war | Pearson Correlation | **-0.071\*** | **-0.068\*** | -0.036 |
|  | P-value | **0.013** | **0.018** | 0.206 |

Interestingly, the number of children who passed away due to starvation has a weak positive correlation with the HFSSM score but not with the other two measures. Weight loss since the war started shows a weak but statistically significant negative correlation with the HFIAS score only. This implies that people who lost more weight tend to have higher scores on the HFIAS, indicating greater food insecurity and the lack of food as the main cause of weight loss, nothing else. Weight before the war did not have significant correlations with any hunger scores, implying that the damaging effect of the Israeli attacks affected all people regardless of their economic, health, or residential status. Current weight and BMI were negatively and significantly correlated with the HHS only, indicating the impactful effect of the war on reducing body weight as a consequence of the lack of food security and widespread hunger. The information from Table 6 suggests that factors like displacement, age, and children’s health may be important when considering hunger and food insecurity in this population.

The cross-tabulation and chi-square analysis of HSS against sociodemographic variables reveal significant findings. The seriousness of household hunger varies significantly with the city (P < 0.001), and the results revealed significant differences in household hunger levels across the three regions. In the Northern region (Gaza City and North Gaza), about a quarter (25.5%) reported varying degrees of hunger, with 130 households experiencing moderate hunger and 178 households facing severe hunger (P < 0.001). To a lesser extent, the Middle region (Deir Al Balah) reported varying degrees of hunger (247 households, about 20%), with about 156 households reporting moderate hunger and 91 reported experiencing severe hunger. Conversely, in the Southern region (Khan Younis and Rafah), 42% (513 households) reported varying degrees of hunger, with 207 facing moderate hunger and a notable 306 households experiencing severe hunger. Overall, the findings suggest a significant variation in household hunger levels across different governorates, with the Southern region exhibiting the highest percentages of severe hunger. There was no significant difference in hunger severity between males and females, implying that sex is not a determinant factor, and the Israeli attacks did not differentiate between the sensitive and non-sensitive, vulnerable and non-vulnerable population groups. However, households where a woman was responsible for the family exhibited higher hunger severity (P< 0.001).

Economic status before the war also played a crucial role (P < 0.001), with households having medium economic status experiencing higher hunger severity. In contrast, expectedly, those with high economic status showed a lesser extent of hunger severity. Expectedly, the appearance of starvation symptoms is significantly associated with higher hunger severity (P < 0.001). Hunger severity was higher in households where the partner was not working (P < 0.001). The educational level of the partner was significantly related to hunger severity (P = 0.003), with lower educational levels (primary and elementary) associated with higher hunger severity, while households with partners having BSc or higher degrees experienced less severe hunger. The condition of the house significantly affects hunger severity (P < 0.001), with higher severity in households with total or partial house destruction (Table 7). The type of address before the war showed significant variation (P = 0.001), with higher hunger severity in households originally from city dwellers and less severe hunger in camps. Current living conditions also play a significant role (P < 0.001), with higher hunger severity in households living in tents and less severe hunger in those living in homes. Lastly, receiving help from relief organizations significantly influences hunger severity (P < 0.001), with higher severity in households receiving intermittent help and less severe hunger in those not receiving or receiving regular help. Overall, the analysis demonstrates that household hunger severity is significantly impacted by various sociodemographic factors, including city, family responsibility, economic status, starvation symptoms, partner’s working status, educational level, house condition, previous and current addresses, and relief organization support (Table 7).

**Table 7:** HSS vs. sociodemographic cross-tabulation and chi-square analysis.

| **Variable** |  | **Little or no household hunger** | **Moderately household hunger** | **Severely household hunger** | **P-value** |
| --- | --- | --- | --- | --- | --- |
| **City** | Northern (Gaza City and North Gaza) | 23 (16.3) | 130 (26.4) | 178 (31.0) | **< 0.001\*** |
|  | Middle (Deir Al Balah) | 48 (34.0) | 156 (31.6) | 91 (15.8) |  |
|  | Southern (Khan Younis and Rafah) | 70 (49.7) | 207 (42.0) | 306 (53.2) |  |
| **Sex** | Male | 65 (46.1) | 227 (46.1) | 270 (46.9) | 0.967 |
|  | Female | 76 (53.9) | 265 (53.9) | 306 (53.1) |  |
| **Responsible for the family** | Man | 131 (92.9) | 462 (93.9) | 494 (85.8) | **< 0.001\*** |
|  | Woman | 10 (7.1) | 30 (6.1) | 82 (14.2) |  |
| **Economic status before the war** | Low | 24 (17.0) | 178 (36.2) | 210 (36.5) | **< 0.001\*** |
|  | Medium | 96 (68.1) | 288 (58.5) | 314 (54.5) |  |
|  | High | 21 (14.9) | 26 (5.3) | 52 (9.0) |  |
| **Starvation symptoms appearance** | Yes | 86 (61.0) | 402 (81.7) | 525 (91.1) | **< 0.001\*** |
|  | No | 55 (39.0) | 90 (18.3) | 51 (8.9) |  |
| **Did any child die because of starvation** | Yes | 0 (0.0) | 2 (0.4) | 3 (0.5) | 0.688 |
|  | No | 141 (100.0) | 490 (99.6) | 573 (99.5) |  |
| **Marital status** | Married | 136 (96.5) | 471 (95.7) | 554 (96.2) | 0.207 |
|  | Widowed | 1 (0.7) | 16 (3.3) | 16 (2.8) |  |
|  | Divorced | 4 (2.8) | 5 (1.0) | 6 (1.0) |  |
| **Partner working** | Yes | 75 (53.2) | 210 (42.7) | 172 (29.9) | **< 0.001\*** |
|  | No | 66 (46.8) | 282 (57.3) | 404 (70.1) |  |
| **The educational level of the partner** | Primary | 2 (1.4) | 4 (0.8) | 29 (5.0) | **0.003\*** |
|  | Elementary | 13 (9.2) | 69 (14.0) | 61 (10.6) |  |
|  | Secondary | 43 (30.5) | 166 (33.7) | 175 (30.4) |  |
|  | BSc | 79 (56.0) | 233 (47.4) | 281 (48.8) |  |
|  | MSc | 3 (2.1) | 16 (3.3) | 27 (4.7) |  |
|  | PhD | 1 (0.7) | 4 (0.8) | 3 (0.5) |  |
| **House status** | Total destruction | 57 (40.4) | 279 (56.7) | 321 (55.7) | **< 0.001\*** |
|  | Partial destruction | 53 (37.6) | 139 (28.3) | 180 (31.3) |  |
|  | No Destruction | 21 (14.9) | 45 (9.1) | 32 (5.6) |  |
|  | I do not know | 10 (7.1) | 29 (5.9) | 43 (7.5) |  |
| **Address before war** | City house/Home | 102 (72.3) | 425 (86.3) | 454 (78.8) | **0.001\*** |
|  | Camp | 39 (27.7) | 67 (13.6) | 122 (21.2) |  |
| **Current address** | Tent | 72 (51.1) | 250 (50.8) | 385 (66.8) | **< 0.001\*** |
|  | Home | 47 (33.3) | 91 (18.5) | 70 (12.2) |  |
|  | School | 22 (15.6) | 151 (30.7) | 121 (21.0) |  |
| **Do you receive any help from a relief organization** | No | 8 (5.7) | 107 (21.7) | 120 (20.8) | **< 0.001\*** |
|  | Regularly | 6 (4.3) | 9 (1.8) | 15 (2.6) |  |
|  | Intermittent | 127 (90.1) | 376 (76.4) | 441 (76.6) |  |

The logistic regression analysis of predictors of food hunger shows significant findings across various sociodemographic variables, which are presented in Table 8. Using Northern Gaza (Gaza City and North Gaza) as the reference, the Middle region is about three times more likely to report hunger (OR, 2.771, 95% CI, 1.273 to 6.030, P = 0.010). The same, but to a lesser extent, for the Southern region (Khan Younis and Rafah) (OR, 1.930, 95% CI, 1.076 to 3.461, P = 0.027). These results indicate that individuals in Deir Al Balah and Khan Younis/Rafah were significantly more likely to experience food insecurity and hunger compared to those in Northern Gaza.

**Table 8:**
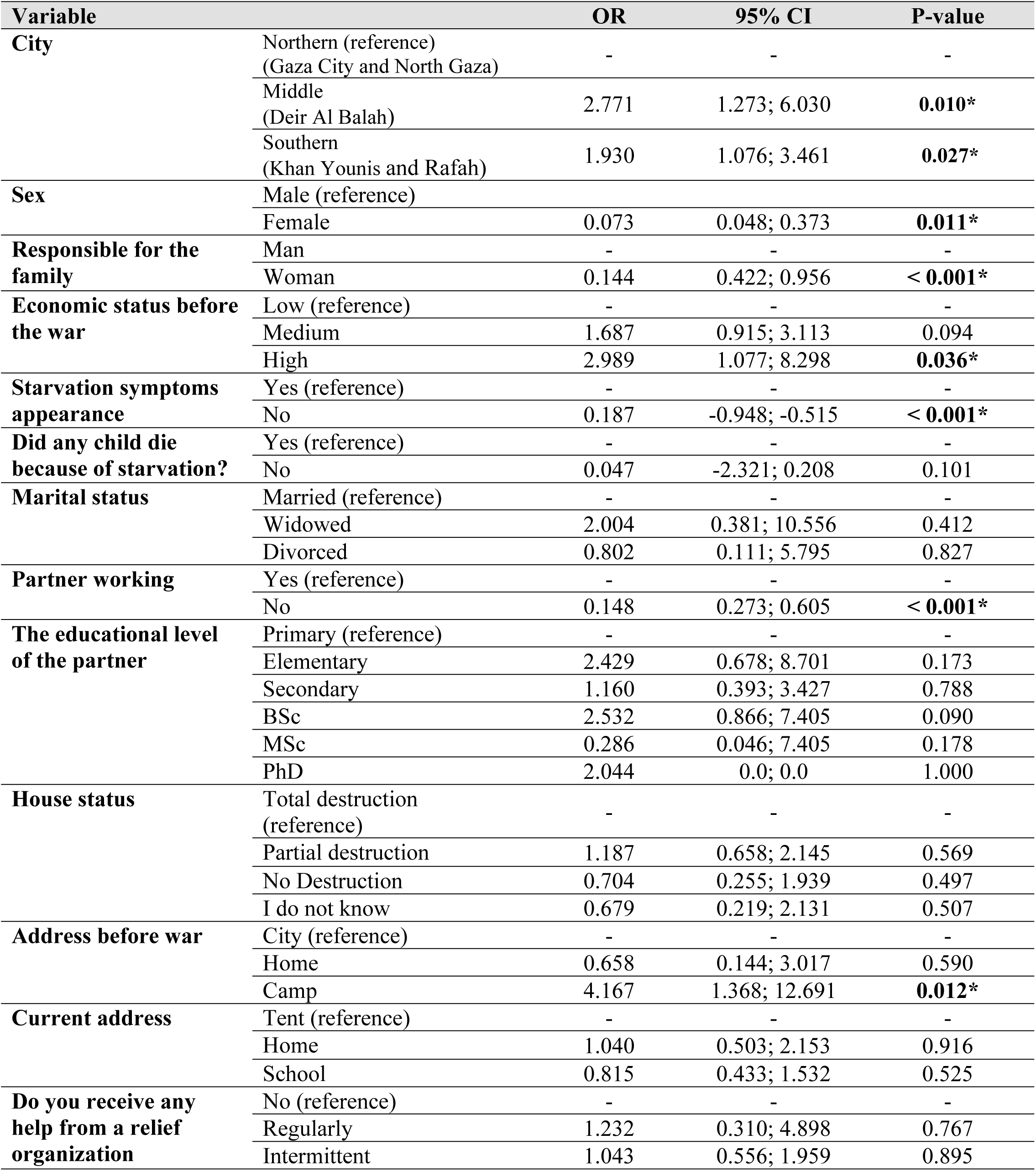
Predictors of food insecurity and hunger.

Sex is an important predictor, with females having lower odds of food hunger (OR = 0.073, 95% CI: 0.048-0.373, P = 0.011) compared to males (reference). Responsibility for the family significantly affects food hunger, with households where a woman is responsible having lower odds (OR = 0.144, 95% CI: 0.422-0.956, P < 0.001). Economic status before the war is also a significant predictor, with high economic status associated with higher odds of food hunger (OR = 2.989, 95% CI: 1.077-8.298, P = 0.036) compared to low economic status (reference). The appearance of starvation symptoms significantly predicts food hunger, with households not experiencing starvation symptoms having lower odds (OR = 0.187, 95% CI: -0.948 - -0.515, P < 0.001) (Table 8).

The partner’s working status is substantial, with households where the partner is not working having lower odds of food hunger (OR = 0.148, 95% CI: 0.273-0.605, P < 0.001). Address before the war is significant, with households from camps having higher odds of food hunger (OR = 4.167, 95% CI: 1.368-12.691, P = 0.012) compared to city residents (reference)(Table 8).

Overall, significant predictors of food hunger include the city of residence, sex, responsibility for the family, economic status before the war, the appearance of starvation symptoms, partner’s working status, and address before the war. At the same time, other variables do not show significant predictive value.

## Discussion

This study represents the first-ever original research work released from Gaza after October 7^th^, and it stands as the first population-based study on the prevalence of hunger in the region following this damaging war on the Strip. By employing quantitative research, this paper seeks to provide a comprehensive understanding of the current food security and hunger situation in the five governorates of the Gaza Strip, contributing to the broader discourse on humanitarian aid and conflict resolution. Significant findings included high levels of food insecurity, with 100% of households experiencing some level of food insecurity according to HFSSM and 97.7% being severely food insecure per HFIAS. The results revealed a catastrophic, unprecedented, extremely high prevalence of hunger, reaching more than 95%. The HHS indicated that 88% of households experienced varying degrees of hunger. The war led to significant weight loss among individuals, with the average weight dropping from 74.6 kg before the war to 64.1 kg.

This community-based cross-sectional study was conducted to assess household hunger during the Israeli war on Gaza 2023-2024. The study’s target population was all the households in the Gaza Strip, Palestine. The Gaza Strip has long been a focal point of geopolitical conflicts and humanitarian crises. Since the mid-20^th^ century, the region has been shaped by complex historical events, including the 1948 Arab-Israeli War, the 1967 Six-Day War, the subsequent Israeli occupation, and the recently aggressive damaging war on Gaza 2023-2024. The blockade imposed by Israel has severely restricted the movement of people and goods, contributing to Gaza’s economic decline. The blockade, combined with recurrent Israeli attacks and violence on Gaza, has devastated Gaza’s infrastructure, crippled its economy, and left the majority of its 2.3 million inhabitants dependent on humanitarian aid. Unemployment rates are among the highest in the world, with youth unemployment exceeding 80%. The local economy is characterized by chronic shortages of basic goods, rampant poverty, and an almost non-existent manufacturing sector. In recent months, the situation in Gaza has deteriorated to an unprecedented level, culminating in a full-blown famine. Several factors have contributed to this crisis. The intensification of the blockade, particularly restrictions on food and fuel imports, has severely disrupted food supply chains. Agricultural production within Gaza has plummeted due to a lack of resources and infrastructure damage, further limiting local food availability. Additionally, recent aggressive wars have displaced almost all families, destroyed homes, and obliterated agricultural lands, exacerbating food insecurity.

The nutritional status of the Palestinian population, particularly in the Gaza Strip and the West Bank, is profoundly impacted by food insecurity, poverty, military occupation, and political instability. Malnutrition, especially in early childhood, poses significant risks for impaired cognitive and physical development and increased susceptibility to infectious diseases. Micronutrient deficiencies, particularly iron, vitamin A, and iodine, are critical issues, with high prevalence rates of iron-deficiency anemia among children and pregnant women [3,14,15,16,17].

Before the 7^th^ October crisis, food insecurity was a major concern, with 60% of households in the Gaza Strip facing moderate to severe food insecurity. The poverty rate in the Gaza Strip was notably high at 53%, compared to 13.9% in the West Bank. One-fifth of children in the Gaza Strip were stunted by the age of two, with girls being more affected than boys. Exclusive breastfeeding rates are below the WHO recommendations, with only 24.4% of infants in the Gaza Strip being exclusively breastfed compared to a national rate of 39.9%. The ongoing military occupation, along with sieges and curfews, significantly hampers food availability and access, contributing to the deteriorating nutritional status of the population. Overcrowded living conditions, particularly in the Gaza Strip, exacerbate the challenges related to nutrition and health, with a population density of 5,138 inhabitants per km², much higher than the West Bank’s 500 inhabitants per km². Addressing malnutrition and nutritional shifts requires multi-sectoral interventions involving healthcare, education, and agriculture. Formulating and implementing evidence-based national nutritional policies can help prioritize interventions and allocate resources effectively to tackle these challenges. These insights underscore the critical need for targeted nutritional interventions and policies to improve the overall health and well-being of the Palestinian population, particularly among the most vulnerable groups such as children and women [17].

The ongoing damaging war on the occupied Strip has exacerbated the already severe food insecurity that the people of Gaza have faced since the blockade was enforced in 2007. Several factors are believed to be associated with the devastating effect on food security and the high prevalence of hunger in the Gaza Strip due to the recent war. These factors include incessant shelling and ground operations that have inflicted significant damage or completely destroyed agricultural land and food production equipment, such as bakeries, mills, and food processing facilities, leading to the collapse of Gaza’s food system. The dire situation has forced several families, particularly in northern Gaza, to consume animal feed and plants to survive. Experts and human rights organizations widely regard Gaza as experiencing the most severe hunger crisis globally, with its population on the brink of famine, if not already experiencing it. The deliberate use of starvation as a tactic in warfare has contributed to this extreme humanitarian catastrophe, which threatens to impact global food security across all six fundamental aspects, endangering the achievement of SDG 2. Although international organizations are currently working to alleviate the severe food shortage and hunger, it is crucial to develop more comprehensive and lasting strategies to address the underlying causes of food insecurity in Gaza. This will help ensure that all inhabitants have access to a sufficient and nourishing diet [3].

In 2022, a report from the Gaza Strip revealed that children in food-insecure homes had a significant occurrence of moderate underweight (30.4%), stunting (32.8%), wasting (9.6%), and acute undernutrition (30.4%). This research was published approximately two years before October 7^th^. Before the damaging war, there were notable disparities in weight, height/length, mid-upper arm circumference, weight-for-age and mid-upper arm circumference z-scores, underweight, acute undernutrition, protein intake, fat intake, vitamin D intake, zinc intake, continued breastfeeding, nutrition-related knowledge, nutrition-related attitudes, and minimum dietary diversity score between the food-insecure and food-secure groups. In addition, approximately 56.0% of households experiencing food insecurity lack sufficient information about nutrition, 77.6% hold negative attitudes towards nutrition, and 95.2% fail to meet the minimum dietary diversity score. Overall, children from households experiencing food insecurity had a significant occurrence of moderate underweight, stunting, wasting, and acute undernutrition. Furthermore, the combination of low socioeconomic status, inadequate dietary intake, insufficient knowledge, attitudes, and practices linked to nutrition, and a lack of variety in the diet all contributed to the elevated levels of food insecurity observed in children under the age of five in the Gaza Strip before the damaging war [14]. However, all these devastating conditions worsened after October 7th.

Before the current situation, more than 75% of the Gaza Strip’s population relied on assistance, as shown in the report of the Global Nutrition Cluster (GNC) Report of February 2024 [15], with reliance on humanitarian aid ranging from 70% to as high as 85% in the different governorates of the Strip. In the GNS report, food insecurity ranged from 60.9% in the Gaza governorate to 69.5% in the Rafah governorate. Food insecurity went in line with the assistance level in the five governorates of the Strip, where the highest rate of reliance on humanitarian assistance was in Rafah (85.1%), down to the lowest rate in Gaza (70.9%)[15].

Assessing the nutritional status of drivers in the Gaza Strip by the GNC report seven months ago showed a devastating situation that is consistent with the catastrophic current situation seen now[15]. The analysis of the different drivers for the nutritional status was as follows: Dietary Diversity in children 6-23 months and Dietary diversity in pregnant and breastfeeding women (PBW) were extremely critical in four out of the five governorates (North Gaza, Gaza City, Deir Al Balah, Khan Younis, and Rafah); Children reporting one or more diseases, Diarrhea (for children under five years, CU5), and other diseases (fever, vomiting, skin infection) were extremely critical in two governorates (Deir Al Balah and Rafah); and finally, water and sanitation access was extremely critical in three governorates (North Gaza, Deir Al Balah, and Rafah). However, acute respiratory infection (ARI) in CU5 was critical in Deir Al Balah and Rafah. These data reveal that Rafah and Deir Al Balah were the governorates most affected by malnutrition in the Gaza Strip[15].

The humanitarian crisis in the Gaza Strip, marked by unprecedented, catastrophic levels of hunger following the actions of Israeli forces after October 7th, 2024, raises significant concerns under international law, human rights frameworks, and the rules governing war crimes. International Humanitarian Law (IHL), specifically the Geneva Conventions and their Additional Protocols, regulates the conduct of armed conflicts and aims to protect individuals who are not participating in hostilities, including civilians [18]. The Fourth Geneva Convention stipulates that civilians should be protected against acts of violence, including starvation, as a method of warfare.

At the same time, Article 54 of Protocol I, in addition to the Geneva Conventions, explicitly prohibits attacking, destroying, removing, or rendering useless objects indispensable to the survival of the civilian population, such as foodstuffs and agricultural areas [19]. Customary international law also emphasizes the protection of civilians in conflict zones and prohibits starvation as a method of warfare. The right to food is a fundamental human right recognized in Article 25 of the Universal Declaration of Human Rights (UDHR) and Article 11 of the International Covenant on Economic, Social, and Cultural Rights (ICESCR). This right entails the availability of food in sufficient quantity and quality to satisfy the dietary needs of individuals, free from adverse substances, and acceptable within a given culture. As the occupying power in Gaza, Israel has obligations under international human rights law to ensure the population’s access to food and not to take actions that would undermine this access. The severe restrictions on the movement of goods and people, which impede access to food and other essentials, could be a violation of these obligations. The ICESCR also recognizes the right to the highest attainable standard of health (Article 12), which is intrinsically linked to the right to food.

Starvation and malnutrition severely impact health, leading to increased morbidity and mortality. The Rome Statute of the International Criminal Court (ICC) defines the intentional starvation of civilians as a war crime under Article 8(2)(b)(xxv), which includes depriving civilians of objects indispensable to their survival, such as food and water. If actions by Israeli forces deliberately lead to starvation or deny food and medical supplies to civilians, these could constitute war crimes. Investigations and prosecutions by the ICC or other international tribunals could be warranted if substantial evidence supports such allegations. Collective punishment, where a group is penalized for the actions of a few, is prohibited under IHL. Indiscriminate measures that lead to widespread suffering among the civilian population of Gaza could be considered collective punishment and, thus, a violation of international law. The catastrophic levels of hunger in the Gaza Strip post-October 7, 2024, have profound implications under international law, human rights norms, and war crimes statutes. Israeli actions that result in widespread deprivation of food and essentials may constitute violations of IHL and human rights obligations, potentially rising to the level of war crimes if starvation is used as a method of warfare. Ensuring accountability and providing humanitarian relief are critical to addressing the severe humanitarian impact on the civilian population in Gaza.

A new report by the Integrated Food Security Phase Classification (IPC) confirms the catastrophic state of food insecurity in Gaza [4]. According to the report, the entire Gaza Strip remains at a high risk of famine due to ongoing war on the occupied Strip and restricted humanitarian access. The report indicates that 96% of the population, or 2.15 million people, are experiencing acute food insecurity (IPC Phase 3 or higher), with 495,000 individuals facing catastrophic levels of food insecurity (IPC Phase 5) until at least September 2024. The severity of the situation underscores the urgent need to ensure that food and other supplies reach all residents in Gaza. The report emphasizes that only a cessation of hostilities combined with sustained humanitarian access can mitigate the risk of famine in the Gaza Strip [4].

Though the current work presents a novel, first-of-its-kind original research work since the beginning of the war in the Gaza Strip after October 7^th^ and covers a large sample size for the households in the occupied Strip, the current work entails a list of limitations that should be considered. While HFIAS offers a focused examination of food access and the severity of food insecurity through nine specific questions, its critics argue that it may oversimplify complex food insecurity experiences, especially in diverse cultural contexts [20]. On the other hand, HFSSM, with its comprehensive 18-question framework, provides a broader, more detailed picture of food security over the past year. However, detractors claim that its extensive nature can lead to respondent fatigue and potential inaccuracies [21]. Both tools, despite their widespread use in research and policy-making, face scrutiny over their effectiveness and adaptability in different settings, highlighting the ongoing debate over the best methods to capture the multifaceted nature of food insecurity and hunger.

## Conclusions

The analysis of sociodemographic variables and their association with household hunger severity and food insecurity highlights a critical situation in Gaza. The high prevalence of food insecurity and hunger, reaching levels of famine, is evident across multiple indicators. The significant predictors of food hunger include city of residence, sex, responsibility for the family, economic status before the war, appearance of starvation symptoms, partner’s working status, and address before the war. The data shows that residents of certain areas, particularly Middle GA and camp dwellers, are more severely affected. Households with women responsible for the family, those with low economic status, and those experiencing starvation symptoms are particularly vulnerable. The findings underscore the dire consequences of the Israeli attacks on the Gaza Strip, leading to widespread destruction of the infrastructure and displacement. The high rates of household hunger severity, the appearance of starvation symptoms, and the significant predictors of food insecurity reflect the urgent need for humanitarian intervention. It is imperative to cease fire and initiate comprehensive relief efforts to address the immediate needs of the affected population and to rebuild the socioeconomic fabric of Gaza. The international community must prioritize the cessation of hostilities and provide sustained support to alleviate the food insecurity and hunger crisis in Gaza.

## Authors’ Contributions

MEF and ASA conceptualized, supervised, and led the study: AMF, RMF, MMA, SAS, SSA, HSB, NAA, NAN, MKA, RMA, KIG, MIA, SSA, AOA, and RAB. ASA, RMF, and NAB contributed to the data collection and data entry; MEF, ANA, NAB, RMF, MOS, and ZGA contributed to the data analysis; MEF and ASA wrote the manuscript. All authors participated in the review of the manuscripts and read and approved the final manuscript.

## Data Availability

All relevant data are within the manuscript and its Supporting Information files.

https://docs.google.com/spreadsheets/d/19oMCAgcjNfF0XM9PHcfiUeUFnS_Vz9Pg/edit?usp=sharing&ouid=103274294772557410736&rtpof=true&sd=true

## Acknowledgments

We would like to convey our gratitude to all of the study participants and the Palestinian Ministry of Health for permitting us to conduct this study.

## Ethics approval and consent to participate

The Palestinian Ministry of Health approved the present study. The participants gave written consent and were informed of their rights to reject or withdraw from the study at any time.

## Consent for publication

Written informed consent for the publication was obtained from the participants.

## Availability of supporting data

All the datasets during and/or analyzed during the current study are available from the corresponding author upon reasonable request.

## Funding

This study did not receive a grant from any institution or organization.

Data repository link: https://docs.google.com/spreadsheets/d/19oMCAgcjNfF0XM9PHcfiUeUFnS_Vz9Pg/edit?usp=sharing&ouid=103274294772557410736&rtpof=true&sd=true

## References

1. Khatib R, McKee M, Yusuf S (2024) Counting the dead in Gaza: difficult but essential. The Lancet.

2. Alaimo K, Chilton M, Jones SJ (2020) Chapter 17 - Food insecurity, hunger, and malnutrition. In: Marriott BP, Birt DF, Stallings VA, Yates AA, editors. Present Knowledge in Nutrition (Eleventh Edition): Academic Press. pp. 311–326.

3. Hassoun A, Al-Muhannadi K, Hassan HF, Hamad A, Khwaldia K, et al. (2024) From acute food insecurity to famine: how the 2023/2024 war on Gaza has dramatically set back sustainable development goal 2 to end hunger. Frontiers in Sustainable Food Systems 8.

4. TIFSPCIFRC (2024) FAMINE REVIEW COMMITTEE: GAZA STRIP, JUNE 2024.

5. Haan N, Hailey P, Maxwell D, Seal A, Lopez J. Famine Review Committee: Gaza Strip, March 2024, Conclusions and Recommendations; 2024. IPC.

6. 6. Deitchler M, Ballard T, Swindale A, Coates J (2011) Introducing a simple measure of household hunger for cross-cultural use.

7. Maxwell D, Adan G, Hailey P, Day M, Odhiambo SB, et al. (2023) Using the household hunger scale to improve analysis and classification of severe food insecurity in famine-risk conditions: Evidence from three countries. Food Policy 118: 102449.

8. Maxwell D, Adan G, Hailey P, Day M, Odhiambo SBJ, et al. (2023) Using the household hunger scale to improve analysis and classification of severe food insecurity in famine-risk conditions: Evidence from three countries. Food Policy 118: 102449.

9. Demie TG, Gessese GT (2023) Household food insecurity and hunger status in Debre Berhan town, Central Ethiopia: Community-based cross-sectional study. Front Nutr 10.

10. Kleve S, Gallegos D, Ashby S, Palermo C, McKechnie R (2018) Preliminary validation and piloting of a comprehensive measure of household food security in Australia. Public Health Nutrition 21: 526–534.

11. Gichunge C, Harris N, Tubei S, Somerset S, Lee P (2015) Relationship between food insecurity, social support, and vegetable intake among resettled African refugees in Queensland, Australia. Journal of hunger & environmental nutrition 10: 379–389.

12. Coates J, Swindale A, Bilinsky P (2007) Household Food Insecurity Access Scale (HFIAS) for measurement of food access: indicator guide: version 3.

13. WHO (2010) A healthy lifestyle - WHO recommendations. World Health Organization.

14. El Bilbeisi AH, Al-Jawaldeh A, Albelbeisi A, Abuzerr S, Elmadfa I, et al. (2022) Households’ Food Insecurity and Their Association With Dietary Intakes, Nutrition-Related Knowledge, Attitudes and Practices Among Under-five Children in Gaza Strip, Palestine. Frontiers in Public Health 10.

15. 15. Cluster GN (2024) Nutrition Vulnerability and Situation Analysis Gaza.

16. Abuzerr S, Al-Jawaldeh A, Ashour Y, Zinszer K, El Bilbeisi AH (2024) The silent crisis: effect of malnutrition and dehydration on children in Gaza during the war. Frontiers in Nutrition 11.

17. A. Assaf E, Al Sabbah H, Al-Jawadleh A (2023) Analysis of the nutritional status in the Palestinian territory: a review study. Frontiers in Nutrition 10.

18. (ICRC) ICotRC (2014) The Geneva Conventions of 1949 and their Additional Protocols.

19. International Humanitarian Law Database nCoRCI (2024) Article 54: Protection of objects indispensable to the survival of the civilian population.

20. Lee JS, Zopluoglu C, Andersen LS, Stanton AM, Magidson JF, et al. (2021) Improving the measurement of food insecurity among people with HIV in South Africa: a psychometric examination. Public Health Nutr 24: 3805–3817.

21. Calloway EE, Carpenter LR, Gargano T, Sharp JL, Yaroch AL (2023) New measures to assess the "Other" three pillars of food security-availability, utilization, and stability. Int J Behav Nutr Phys Act 20: 023–01451.

